# Deep learning models to predict mammographic density jointly on standard dose and low dose images

**DOI:** 10.1101/2024.04.10.24305572

**Authors:** Steven Squires, Alistair Mackenzie, D. Gareth Evans, Sacha J Howell, Susan M Astley

**Affiliations:** University of Exeter, Exeter, United Kingdom; NCCPM, Royal Surrey NHS Foundation Trust, Guildford, United Kingdom; University of Manchester, Manchester, United Kingdom

## Abstract

**Objectives:** Mammographic density is associated with increased risk of developing breast cancer. Automated estimation of density in women below normal screening age would enable earlier risk stratification. We are piloting the use of low dose mammograms combined with models that can make accurate mammographic density estimates.

**Methods:** Three models were trained on a joint set (107,619) of standard dose mammograms with associated density scores and their simulated low dose counterparts such that the models made predictions on standard and low dose mammograms. A second set of models was trained separately on the standard and simulated low dose mammograms. All models were tested on a held-out set from the training data and an independent dataset with 294 pairs of standard and real low dose mammograms.

**Results:** The root mean squared errors (RMSE) between the model predictions and density scores on standard and simulated low dose images were 8.26 (8.16-8.36) and 8.27 (8.17-8.38) respectively. The RMSE between predictions on standard and simulated low dose images for the jointly trained models was 1.91 (1.88-1.96). The RMSE of the predictions on the real low dose images compared to the standard dose images is 3.79 (2.75-4.99).

**Conclusions:** Deep learning models make density predictions on low dose images with similar quality as on standard dose images. Such automated analysis of low dose mammograms could contribute to accurate breast cancer risk estimation in younger women enabling stratification for further monitoring and preventative therapy.

**Advances in knowledge:** Mammographic density can be estimated in low dose mammograms with similar quality to standard dose mammograms.

## 1) Introduction

High breast density, usually defined as the relative proportion of radio-opaque fibroglandular to radiolucent fatty tissue in the breast, is known to be a risk factor for developing cancer (1) and is also associated with a decreased ability to detect breast cancers mammographically due to masking (2). Whilst screening usually starts at around age 50 in the United Kingdom, around one in five breast cancers are detected at an earlier age. Producing accurate mammographic density predictions for women below screening age could enable personalisation of screening with better targeting of alternative or supplementary imaging modalities or changes to the frequency of screening. However, full field digital mammography, in which radiologists’ estimates of density from mammograms show a particularly strong relationship with cancer risk (3), is not recommended for younger women because of the risk of radiation induction of tumours and the increased density resulting in more frequent recalls. An alternative is to utilise mammography with substantially reduced radiation dose with automated density analysis. An ongoing project aims to assess ways of predicting breast cancer risk in women aged 30-39 including the use of low dose mammography (4). The aim of this paper is to investigate how to produce automated mammographic density predictions on low dose images that are comparable in quality to predictions on standard dose images.

Previous work on density estimation in low dose mammography has primarily utilised three strategies (5). One is to use models trained on standard dose mammograms (6) for direct inference on low dose images with the addition of small alterations to the output to correct for systematic differences. The second approach is to train models on simulated low dose mammograms and use those models for low dose prediction (7,8). The third is to use the low dose mammograms to fine tune a model pre-trained on standard dose images (5).

The first approach utilises models trained on standard dose mammograms and makes direct predictions on low dose images. This assumes a high enough degree of similarity between the low and standard dose images that any differences in density predictions are small or can be corrected post hoc, which has been partially shown in previous work (5). This may be due to the image features relating to mammographic density being clear enough such that they are not fully obscured in the low dose images where the noise levels are higher, i.e. the density signal in the mammograms is strong, something which has been demonstrated in previous work on density estimation from mammograms (9).

The second approach is to train models on standard dose and simulated low dose images separately. However, the way to assess the quality of low dose predictions is to compare to standard dose predictions; because there are two trained models it is hard to disentangle the effects of different training models and of different image doses. Another problem is that there is likely less information available in the low dose images than standard dose images due to increased noise. Ideally, we would train a model that is able to compensate for that information loss by implicitly filling in the missing information.

The third approach is to fine tune a model on real (rather than simulated) low dose images; this requires a substantial amount of training data. In medical imaging access to such data, especially labelled data, is often a challenge, for example in this study we have access to 147 sets of low dose images. Another problem is “model forgetting” (10) where training on a new task causes the model to perform worse on the previous task, as it can become too well tuned to the new domain. When fine-tuning the model on the small number of low dose images the model may then “forget” about images in the broader image-space and perform worse on low dose images it has not seen than if it had not been fine-tuned on the low dose images.

The approach we set out in this paper is to build a joint model trained on both standard and simulated low dose images together. We believe this approach solves all the previously mentioned issues and we demonstrate that the results produced by our models show both state-of-the-art performance on the standard dose and simulated low dose data as well as showing similar predictions on the real low dose images compared to their standard dose equivalents. The model has been trained on (simulated) low dose images so should be able to correct for any variations between the two dose levels. As there is only one model trained which can make both standard and low dose image predictions the final comparison is not confounded by between model variation. Further, as the model sees both the standard and simulated low dose matched images it should be able to learn, if it is possible to do so, how to compensate for any lost information in the simulated low dose images.

In addition to the jointly trained models, we also train models individually on the standard and simulated low dose images to ensure the jointly trained models are not showing a reduction in performance. We train three separate deep learning model architectures to demonstrate that the results we have are not specific to one model type. Finally, we investigate whether the similarity in model predictions between real low dose and standard dose is the same as between the simulated low dose and standard dose images.

## 2) Methods and materials

### 2.1) Data

We utilise two separate datasets, the first is constructed with mammograms from the Predicting Risk Of Cancer at Screening (PROCAS) study (11). We use this dataset to produce simulated low dose images from physics-based software (12,13). These simulated low dose images are equivalent to the standard dose images except for the simulated alteration in dose. In total we have 107,619 images which we partition, at random, into training (70,171 images), validation (17,605 images) and test sets (19,843 images) always keeping images from one woman within the same partition. During the PROCAS study these images were viewed by two expert readers drawn from a pool who each provided a density score on a visual analogue scale (VAS) between 0 and 100%. The final VAS density score for each mammographic image is the average of the two readers’ scores, which is used as the label for training our models.

The second dataset is from the Automated Low Dose Risk Assessment Mammography (ALDRAM) study (5); this consists of images from 147 women between ages of 30 and 45 who had had cancer in one breast and were attending routine screening. There are standard dose images taken at the four standard mammographic views (right and left breast with craniocaudal and mediolateral oblique views) and whilst the right breast was in compression the dose was reduced to 10% of the standard dose (or the machine minimum) and a second image was taken. The low dose images are thus available for right craniocaudal (RCC) and right mediolateral oblique (RMLO) views and should be directly comparable with the standard dose equivalents. There are no associated VAS density scores with the ALDRAM data.

All images in this study are in the “for-processing” (“raw”) state and are reduced in size down to 640 by 512 pixels whilst maintaining their aspect ratio. Histogram equalisation is applied, and the images are normalised. The same procedure is applied for standard, simulated low dose and low dose images. This is the same procedure applied previously for mammographic density predictions (6,7).

Our aim is to produce a model that can accurately predict mammographic density on low dose images. We have no VAS scores for the ALDRAM dataset (the images have not been scored by expert readers) so accurate predictions on the low dose images are considered with reference to the predictions on the corresponding standard dose images, i.e. the predictions on the standard dose images effectively become the labels. We use both root mean squared error (RMSE) and the Spearman rank correlation coefficient between predictions on standard and low dose images to measure the quality of the model predictions on the low dose images. This also means our models need to accurately estimate mammographic density in standard dose images and simulated low dose images in the PROCAS dataset which can be compared to the VAS density scores.

### 2.2 Joint and independent training

Our approach is to use the training and validation sets from the PROCAS data and train models on a joint dataset of standard and simulated low dose images. The trained model would then be used for inference on standard and low (simulated and real) dose images. As a direct comparison we also train models separately on the standard and simulated low dose image sets. The standard dose image trained models are then tested on just the standard dose images from the test sets. The simulated low dose image trained models are tested on the low dose (simulated and real) images from the test sets.

### 2.3 Models

There are many deep learning model architectures and they are likely to show modest differences in performance. We therefore take a pragmatic approach trading off time and computational requirements with final model performance. We train three separate types of models: ResNet-18 (14), ResNet-50 (14) and DenseNet-161 (15). ResNet-18 has been used before to make mammographic density predictions and has shown good performance (16). The logic of using ResNet-50 is to test the hypothesis that more layers within the same model architecture would improve model performance. We include DenseNet-161 as a separate model from a different family to establish whether the ResNet family has weaknesses on this dataset and problem domain.

All results shown are on the held-out test set from the PROCAS dataset or the independent (not trained on) ALDRAM dataset. Spearman’s rank correlation coefficients and RMSE are shown. The uncertainties are generated by bootstrapping and reported at the 95% level.

### 2.4 Model training

We train the three different model types (ResNet-18, ResNet-50 and DenseNet-161) and the joint/independent approaches with the same procedure where possible to ensure a fair comparison can be made. The models are initialised with weights pre-trained from ImageNet (17). We train with the Adam optimiser (18) with a mean-squared error (MSE) objective function altering only the learning rates. Both ResNet models are trained with a batch-size of 20 while the DenseNet models utilise a batch-size of 5 due to limitations of computer memory. The model parameters are saved at the end of each training epoch if the validation MSE is lower than any previous epoch. The final models are selected using the lowest value of the mean squared error of the validation set – for the joint models this is the combined standard and simulated low dose predictions.

## 3) Results

The metrics of density predictions compared to VAS density scores of the joint models on the PROCAS standard and simulated low dose images are shown in Table 1. Comparable scores for previous work (6) have RMSE of 9.18 and Spearman rank correlation coefficient of 0.798 on the standard dose images. While the results are shown separately for the standard dose and simulated low dose images the predictions are produced by the same model.

**Table 1.**
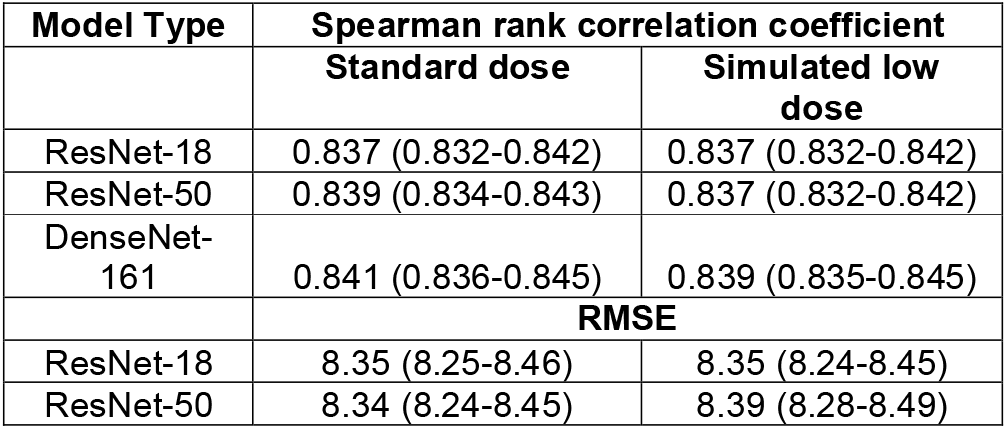
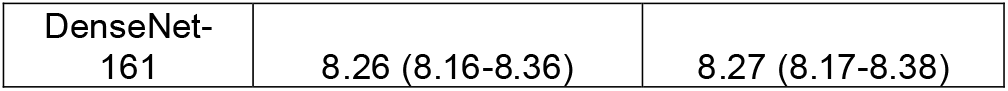
The performance of the three joint models against labels for standard dose and simulated low dose from the PROCAS test set. Results on the standard and simulated low dose images are shown separately but the predictions are produced by the same model.

| Model Type | Spearman rank correlation coefficient |  |
| --- | --- | --- |
|  | Standard dose | Simulated low dose |
| ResNet-18 | 0.837 (0.832-0.842) | 0.837 (0.832-0.842) |
| ResNet-50 | 0.839 (0.834-0.843) | 0.837 (0.832-0.842) |
| DenseNet-161 | 0.841 (0.836-0.845) | 0.839 (0.835-0.845) |
|  | RMSE |  |
| ResNet-18 | 8.35 (8.25-8.46) | 8.35 (8.24-8.45) |
| ResNet-50 | 8.34 (8.24-8.45) | 8.39 (8.28-8.49) |
| DenseNet-161 | 8.26 (8.16-8.36) | 8.27 (8.17-8.38) |

The equivalent results of predictions compared to VAS density scores for the separate models are shown in Table 2. Unlike for the joint models, here the predictions on the standard dose and simulated low dose images are from the two separately trained models, so the table shows results from a total of six models.

**Table 2.** The performance of the six independent models against labels for standard dose and simulated low dose from the PROCAS test set. Results on the standard and simulated low dose images are produced by different models.

| Model Type | Spearman rank correlation coefficient |  |
| --- | --- | --- |
|  | Standard dose | Simulated low dose |
| ResNet-18 | 0.840 (0.835-0.845) | 0.838 (0.832-0.842) |
| ResNet-50 | 0.840 (0.836-0.845) | 0.838 (0.833-0.843) |
| DenseNet-161 | 0.844 (0.839-0.849) | 0.842 (0.837-0.847) |
|  | **RMSE** |  |
| ResNet-18 | 8.28 (8.18-8.38) | 8.40 (8.3-8.5) |
| ResNet-50 | 8.26 (8.16-8.36) | 8.36 (8.26-8.46) |
| DenseNet-161 | 8.24 (8.14-8.33) | 8.30 (8.20-8.40) |

In Table 3 we show metrics to compare predictions on the standard dose images and predictions on the simulated low dose images for the PROCAS test set. Effectively we are considering the predictions on the standard dose models to be the label to test the quality of the predictions on the simulated low dose images. Results for both the joint and individual models are shown in the same table. The joint predictions are generated by the same model while the individual predictions are from the two separately trained models. The joint models produce more similar predictions on standard and simulated low dose images than the individual models.

**Table 3.** The metrics between predictions made on the standard dose images compared to predictions made on the simulated low dose images from the PROCAS dataset. Results from both the joint models and the individual model are shown in the same table. The joint model shows the predictions on the standard and simulated low dose images from one model while the predictions from the individual models are from two separately trained models.

| Model Type | Spearman rank correlation coefficient |  |
| --- | --- | --- |
|  | Joint | Individual |
| ResNet-18 | 0.990 (0.990-0.991) | 0.980 (0.979-0.980) |
| ResNet-50 | 0.989 (0.988-0.989) | 0.979 (0.979-0.980) |
| DenseNet-161 | 0.989 (0.989-0.990) | 0.987 (0.987-0.988) |
|  | **RMSE** |  |
| ResNet-18 | 1.86 (1.82-1.91) | 2.86 (2.80-2.92) |
| ResNet-50 | 2.06 (2.02-2.10) | 2.82 (2.77-2.87) |
| DenseNet-161 | 1.91 (1.88-1.96) | 2.70 (2.66-2.74) |

To ensure the metrics we show do not disguise differences in prediction, in Figure 1 we show plots of the predictions of our final joint models on the PROCAS standard and simulated low dose data for a direct comparison of the models. We show only 2,000, randomly selected, data-points in the plots to improve readability.

**Figure 1.**
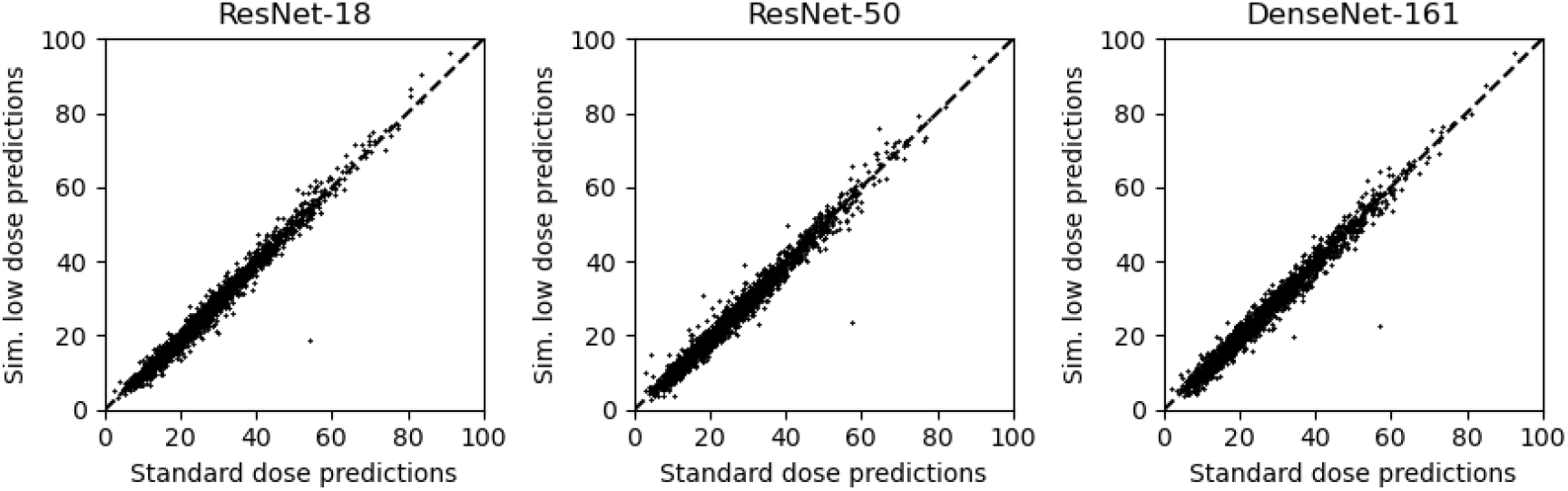
Plots of simulated low dose predictions versus standard dose predictions on the PROCAS data from the three jointly trained models. The number of data-points shown is reduced to 2,000, by random selection, to improve readability.

In Table 4 we present results comparing predictions on the ALDRAM standard and low dose images for both jointly trained models and the individual trained models. The similarity in predictions is reduced compared to the PROCAS results (Table 3). The gap in performance between the joint and individual models has also reduced.

**Table 4.** The metrics between predictions made on the standard dose images compared to low dose images from the ALDRAM dataset.

| Model Type | Spearman rank correlation coefficient |  |
| --- | --- | --- |
|  | Joint | Individual |
| ResNet-18 | 0.980 (0.959-0.991) | 0.979 (0.972-0.983) |
| ResNet-50 | 0.980 (0.969-0.986) | 0.981 (0.975-0.984) |
| DenseNet-161 | 0.978 (0.960-0.989) | 0.986 (0.981-0.989) |
| RMSE |  |  |
| ResNet-18 | 3.72 (2.52-5.00) | 4.19 (3.64-4.78) |
| ResNet-50 | 3.62 (2.91-4.39) | 3.74 (3.37-4.11) |
| DenseNet-161 | 3.79 (2.75-4.99) | 4.41 (3.88-4.92) |

In Figure 2 we show plots of the predictions of the joint models on the ALDRAM standard and low dose data. These are equivalent plots, with predictions from the same models, as in Figure 1.

**Figure 2.**
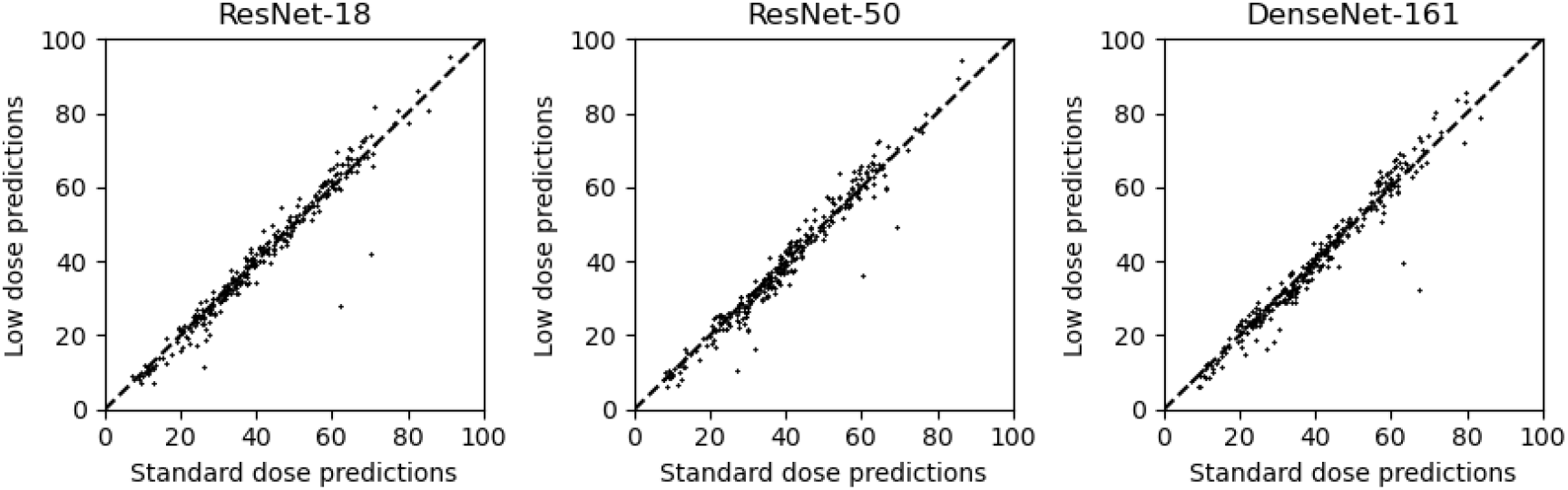
Plots of low dose versus standard dose predictions on the ALDRAM data for the joint models.

In Figure 3 we show how the RMSE between the standard and low (simulated or real) dose joint model predictions changes at different values of the standard dose predictions. We use the predictions on the standard dose images to bin the images into bins of width 30 with a sliding window with stride length of one. Therefore, each value on the y-axis is the RMSE between standard and low dose predictions for those images with a standard dose prediction within 15 points either side of the value on the x-axis. Results are shown for both the PROCAS and ALDRAM datasets. The discontinuities in the ALDRAM results are caused by the small number of data-points in some bins.

**Figure 3.**
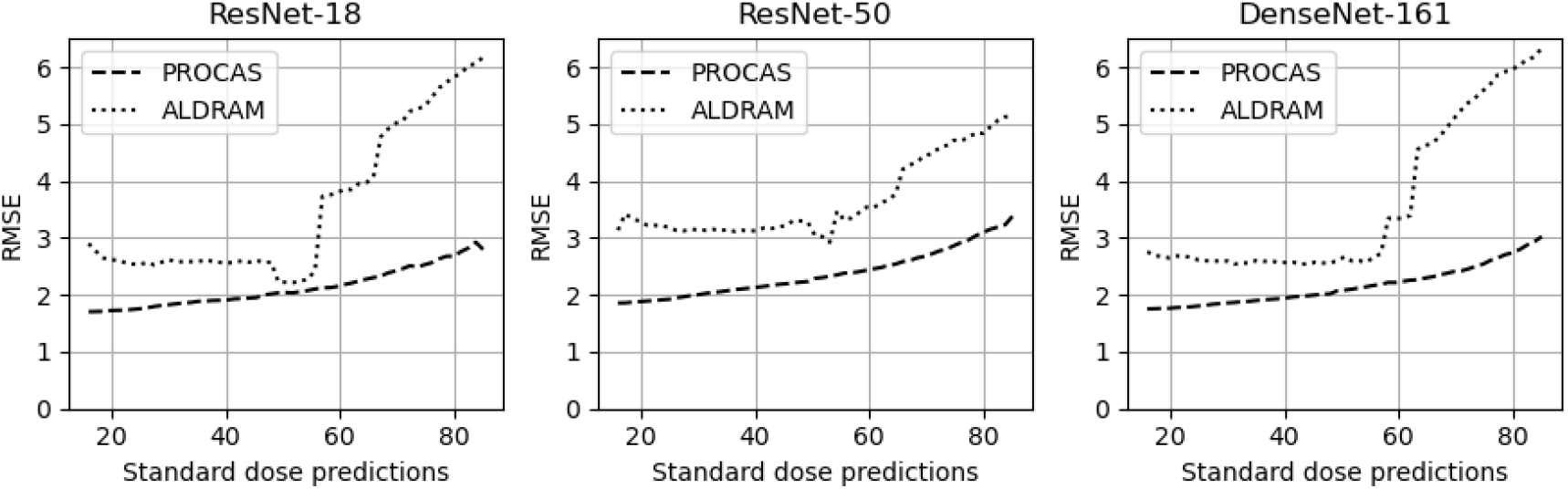
Plots of how the RMSE between standard and low dose predictions changes with the value of the standard dose predictions for both the PROCAS and ALDRAM datasets.

## 4) Discussion

We have shown that our joint models can make accurate predictions on both standard and simulated low dose images; there is no statistically significant reduction in similarity to radiologist scores compared to models trained separately on the two different sets of images. We do not see a significant improvement for the simulated low dose images when training jointly, which implies that including standard dose images alongside simulated low dose images does not improve performance on the simulated low dose images. However, the difference between performance on the standard and simulated low dose images is also statistically insignificant which suggests that the performance gap is small.

The separately trained models perform worse than the jointly trained models when making a direct comparison between standard dose and simulated low dose predictions. This is likely to be due to the removal of model variability with the joint models. This effect is an important advantage of the joint approach as we can now directly compare the performance of the models on the different doses without having to consider how the differences depend on the training of the models.

There is a greater difference in model performance between predictions on the standard and low dose images from the ALDRAM dataset than from the PROCAS dataset (with simulated low dose images). The jointly trained models show this difference clearly with differences in performance between the PROCAS and ALDRAM data. The individual models show these differences too but some of the differences will be due to the different training of the two models making the result less clear.

Automated VAS density prediction models have shown poorer performance at higher densities (6,16). Therefore, part of the reason for the larger model prediction differences on the standard and low dose images in ALDRAM compared to PROCAS is that the ALDRAM images have higher distributions of densities. However, this variation in distribution of densities does not account for all the differences in model similarity as shown in Figure 3. The models perform worse (larger differences between low and standard dose predictions) for all density levels. It is possible that there are differences between the simulated low dose images from PROCAS and the low dose images from ALDRAM which cause the differences in model performance. It is notable that there are outlier points in the ALDRAM plots (Figure 2) which we do not see in the PROCAS plots, even though we show significantly more data for PROCAS. Further research would be valuable to ascertain what is causing these differences and what approaches could improve the results. However, the results (see Figure 2) are very similar with most differences being small between predictions on the standard and low dose images.

## 5) Conclusions

The three deep learning models investigated (ResNet-18, ResNet-50 and DenseNet-161) provide similar density prediction performance on simulated low dose image as on standard dose images for both joint and independently trained models. The jointly trained models show greater similarity in prediction between simulated low dose images and standard dose equivalents compared to separately trained models.

There is a reduction in similarity of predictions between ALDRAM standard and low dose images when compared to PROCAS standard and simulated low dose images. It would be valuable to understand the cause of these differences and, if possible, to improve the models such that they perform as well on the ALDRAM low dose data as they do on the PROCAS simulated low dose data. However, for most images, the predictions between standard and low dose images in the ALDRAM data have only small differences which are unlikely to be problematic when the methods are used clinically. Our work shows that with low dose mammography, automated density assessments can be provided for younger women. This conclusion holds for both jointly trained models and models trained on just simulated low dose images as well as the three model types (ResNet-18, ResNe50 and DenseNet-161) all of which show high performance.

## Data Availability

Data is not currently available to researchers outside of the University of Manchester.

## Notes

### Competing Interest Statement

The authors have declared no competing interest.

### Clinical Trial

ISRCTN55983830

### Funding Statement

The ALDRAM study was funded by the Medical Research Council (UK).

### Author Declarations

Ethics approval for the PROCAS study was through the North Manchester Research Ethics Committee (09/H1008/81). Informed consent was obtained from all participants on entry to the PROCAS study. Ethics approval for the ALDRAM study was granted on 12/02/2019 by the North West - Preston Research Ethics Committee (Barlow House, 3rd Floor, 4 Minshull Street, Manchester M1 3DZ; +44 (0)2071048234; preston.rec{at}hra.nhs.uk), ref: 19/NW/0037

